# High concordance of 70-gene recurrence risk signature and 80-gene molecular subtyping signature between core needle biopsy and surgical resection specimens in early-stage breast cancer

**DOI:** 10.1101/2021.05.28.21257887

**Authors:** Jennifer A. Crozier, Julie Barone, Pat Whitworth, Abraham Cheong, Robert Maganini, Jeffrey Falk, Jia-Perng Wei, Sammy Mee, Jake Ruby, Suoyi Yang, Yen Huynh, Anke Witteveen, Christine Finn, Kate Corcoran, Christa Dreezen, Patricia Dauer, Andrea Menicucci, Shiyu Wang, Annie Tran, Erin Yoder, Bastiaan van der Baan, William Audeh, Annuska Glas, FLEX Investigators Group

## Abstract

**Introduction:** With an increase in neoadjuvant therapy recommendations for most early-stage breast cancer patients due to the COVID-19 pandemic, it has become increasingly imperative to ensure that molecular diagnostic assays provide reliable results from preoperative core needle biopsies. Therefore, the objective of this study was to determine the concordance of MammaPrint results (70-gene signature) and BluePrint results (80-gene signature) between core needle biopsies (CNB) and surgical resection (SR) specimens using prospectively collected matched tissues from patients enrolled in the FLEX trial (NCT03053193).

**Methods:** We analyzed 113 matched CNB and SR tumor specimens from women with early-stage breast cancer enrolled in the FLEX trial. Each patient enrolled in the trial receives a MammaPrint recurrence risk classification test with or without BluePrint molecular subtyping. Concordance of MammaPrint is reported using overall percentage agreement, positive predictive value (PPV, High Risk), negative predictive value (NPV, Low Risk), and Cohen’s kappa coefficient. Additionally, correlations between sample types are reported using Pearson correlation coefficient.

**Results:** We found good concordance for MammaPrint results between CNB and SR tumor samples (90.3%, κ = 0.803), with a 95.1% NPV and 84.6% PPV. There was also a strong correlation of MammaPrint indices between CNB and SR specimens (r = 0.94). In addition to our primary objective, we determined the concordance of BluePrint subtyping in the matched tumor samples, and found excellent concordance (98.2%) and strong correlation of BluePrint scores within each subtype.

**Conclusion:** CNB samples demonstrated overall high concordance with paired SR samples for MammaPrint risk classification, ensuring that physicians are provided with accurate prognostic information for therapy decisions based on testing of core biopsy tissue. Further, BluePrint molecular subtyping also had good concordance between the sample types, outperforming concordance rates based on traditional IHC based classification. Overall, with an increase in neoadjuvant therapy, physicans and patients can be assured that MammaPrint and BluePrint provide reliable results that guide timely and appropriate therapies using preoperative CNB specimens.

## Introduction

Due to the extraordinary circumstances that the COVID-19 pandemic presented last year, health experts and organizations have published treatment guidelines to conserve supplies, triage cases, and reduce risk without compromising patient care^1-4^. As a result, neoadjuvant chemotherapy has become more prevalent in breast cancer treatment plans. Increased neoadjuvant therapy subsequently requires any prognostic information to be obtained from a core needle biopsy (CNB) prior to preoperative chemotherapy.

Multigene assays, like MammaPrint can provide prognostic information which can guide physicians on therapy decisions for patients. However, multigene assays have typically been developed and tested using primary surgical specimens. Only a few studies have determined the performance of multigene assays using CNB in recent years^5-7^. In a large unpaired study, Jakubowski et al. found a similar range of Oncotype Dx Recurrence Scores between CNB and surgical resection (SR) samples (10-22 versus 11-22, respectively)^5^. In two smaller studies using matched CNB and SR samples, researchers reported Pearson correlation coefficients ranging from 0.20-0.99 and overall concordance ranging from 83.9-95%^6,7^.

MammaPrint is a 70-gene assay that can be used to predict the likelihood of recurrence, and response to chemotherapy^8,9^. For example, patients identified as MammaPrint Low Risk can forego chemotherapy without compromising outcome, compared to those identified as High Risk for whom chemotherapy is recommended^9^. We previously demonstrated MammaPrint precision (99.0%) and reproducibility (98.9%) in fresh frozen tissue, with 95% agreement between two sample sites from the same tumor^10^. However, the use of formalin-fixed paraffin -embedded (FFPE) tissue increases multigene assay utility. When we compared matched fresh frozen tissue to FFPE, we demonstrated a concordance of 91.5% in MammaPrint classification and a 0.92 Pearson correlation of MammaPrint indices between samples^11^.

In addition to risk classification, multigene assays (*i*.*e*. BluePrint) can be utilized to reliably subtype women with early-stage breast cancer^12^. We have previously tested the performance of BluePrint molecular subtyping in comparison with immunohistochemistry (IHC) using multiple cohorts and have demonstrated overall subtype reclassification of up to 30%^13-15^. Additionally, tumors reclassified by MammaPrint and BluePrint exhibited more accurate pathological complete response (pCR) rates compared to pCR rates based on their respective clinical subtype^12,14^.

A few small studies have demonstrated some degree of concordance between CNB and SR specimens for their respective multigene assays, but it is unclear how well MammaPrint and BluePrint will perform. In our current study, we have prospectively collected 113 matched CNB and SR specimens from women with confirmed early-stage breast cancer enrolled in the ongoing FLEX study (NCT03053193). The objective of our study is to determine the concordance of MammaPrint and BluePrint results between CNB and SR, to ensure reliable prognostic information can be captured from a CNB.

## Methods

### Patient cohort

FLEX is an ongoing, multi-institutional prospective study of patients with stage I-III early breast cancer. The protocol was approved by Institutional Review Boards at all participating sites and registered with ClinicalTrials.gov (NCT03053193). Enrolled patients consent to a MammaPrint test with/out BluePrint molecular subtyping in addition to clinically annotated gene expression data collection. Patients within the FLEX study (n = 139) with matched CNB and SR specimens were prospectively collected from six institutions. We excluded 26 patients due to neoadjuvant treatment, bilateral or multifocal tumors, prior cancer, borderline MammaPrint CNB results, and/or greater than 90 days from CNB to SR upon final pathology. A total of 113 matched CNB and SR tissues were eligible for this study and passed quality checks.

### Molecular classification

MammaPrint and BluePrint are gene expression diagnostic arrays that use CNB or SR tissue to identify Low Risk versus High Risk tumors, and genomic subtype, respectively^9,12,16^. CNB specimens have been successfully used for MammaPrint and BluePrint in clinical trials for I-SPY2 (NCT01042379) and NBRST (NCT01479101). For this study, patients were identified in our FLEX patient database by their CNB MammaPrint result, and corresponding SR tissue was requested from their respective local institution. Upon receipt of SR FFPE tissue blocks at Agendia (Irvine, CA, USA), sections are prepared. In accordance with diagnostic quality controls and standards, one section per sample was reviewed internally to verify > 30% tumor cellularity. RNA is then isolated with the RNeasy FFPE kit (Qiagen), and concentration measured with the NanoDrop 2000 (Thermo Scientific). cDNA is labeled and hybridized to 44k arrays and scanned using a dual laser scanner (Agilent Technologies) as previously described^17,18^.

MammaPrint Low Risk tumors have a MammaPrint index of > 0.000, and ≤ 0.000 for High Risk tumors. BluePrint classifies tumors as Luminal, Basal, or human epidermal growth factor receptor 2 (HER2) subtype. Together, MammaPrint with BluePrint stratify Luminal subtype tumors into Luminal A (MammaPrint Low Risk) or Luminal B (MammaPrint High Risk). For this study, borderline samples (MammaPrint index between -0.05 and 0.05) were excluded.

### Statistical analysis

Descriptive statistics were used to summarize patient clinicopathological characteristics. To evaluate MammaPrint concordance of High Risk and Low Risk between CNB and SR, overall percentage agreement, positive predictive value (PPV, High Risk), negative predictive value (NPV, Low Risk), and Cohen’s kappa coefficient (*k*) were calculated. Pearson correlation coefficient (r) and Pearson correlation test were calculated using the MammaPrint index for CNB and SR specimens. A 2-sided p-value less than 0.05 is considered statistically significant. Statistical analyses were performed using GraphPad Prism (version 9.0.2) and R (version 4.0.5).

## Results

### Patient Characteristics

A total of 113 patients from the FLEX study database with diagnostic MammaPrint and BluePrint results with matched CNB and SR specimens were included in this study. **Table 1** summarizes clinicopathological characteristics with available data, but excludes data marked as unknown. The majority of patients were over the age of 50 (91/111; 82.0%). 103 of 110 patients (93.6%) were hormone receptor (HR) positive, 7/110 (6.4%) were HR negative, 8/104 (7.5%) were HER2 positive, and 96/104 (89.7%) were HER2 negative. Of the 107 patients with clinical subtyping data, a majority of patients (92/107; 86.0%) had HR positive/HER2 negative tumors, 8 (7.5% had HR positive/HER2 positive tumors, and 7 (6.5% had triple negative (TN tumors. Patients with tumors grade 1-3 (G1-3 were included and out of 108, 42 (38.9% tumors were low grade (G1, 43 (39.8% were intermediate grade (G2, and 23 (21.3% were high grade (G3.

**Table 1:** Patient/tumor clinical characteristics summary

|  | Patient number |
| --- | --- |
| Age<br>(n=2 unknown) |  |
| > 50 | 91 (82.0%) |
| ≤ 50 | 20 (18.0%) |
| Hormone Receptor Status<br>(n=3 unknown) |  |
| HR Positive | 103 (93.6%) |
| HR Negative | 7 (6.4%) |
| HER2 Receptor Status<br>(n=6 unknown) |  |
| HER2 Positive | 8 (7.5%) |
| HER2 Negative | 96 (89.7%) |
| HER2 Equivocal | 3 (2.8%) |
| Clinical Subtype<br>(n=6 unknown) |  |
| HR positive/HER2 negative | 92 (86%) |
| HR positive/HER2 positive | 8 (7.5%) |
| Triple Negative | 7 (6.5%) |
| Tumor Grade<br>(n=5 unknown) |  |
| G1 Low Grade | 42 (38.9%) |
| G2 Intermediate Grade | 43 (39.8%) |
| G3 High Grade | 23 (21.3%) |

### Concordance of MammaPrint results between CNB and SR tumor specimens

The primary planned analysis in this study is the concordance of MammaPrint results between CNB and SR specimens, which were measured using NPV, PPV, overall percentage agreement, and Cohen’s kappa. CNB and SR specimens matched in 44 High Risk and 58 Low Risk patient tumors, resulting in 90.3% overall agreement (κ = 0.803), 95.1% NPV, and 84.6% PPV (**Table 2**. Out of the discordant samples, eight were High Risk on CNB and Low Risk on SR, whereas three were Low Risk on CNB and High Risk on SR. Next, a Pearson correlation test of MammaPrint indices between 113 CNB and SR specimens was performed and resulted in a strong correlation of r = 0.94 (p<0.001 (**Figure 1**.

**Table 2:**
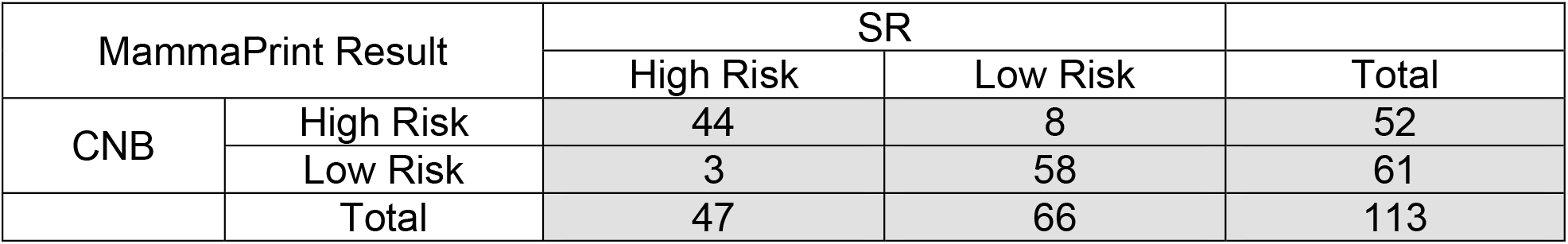
Concordance of MammaPrint results between CNB and SR

| MammaPrint Result |  | SR |  | Total |
| --- | --- | --- | --- | --- |
|  |  | High Risk | Low Risk |  |
| CNB | High Risk | 44 | 8 | 52 |
|  | Low Risk | 3 | 58 | 61 |
|  | Total | 47 | 66 | 113 |

**Figure 1.**
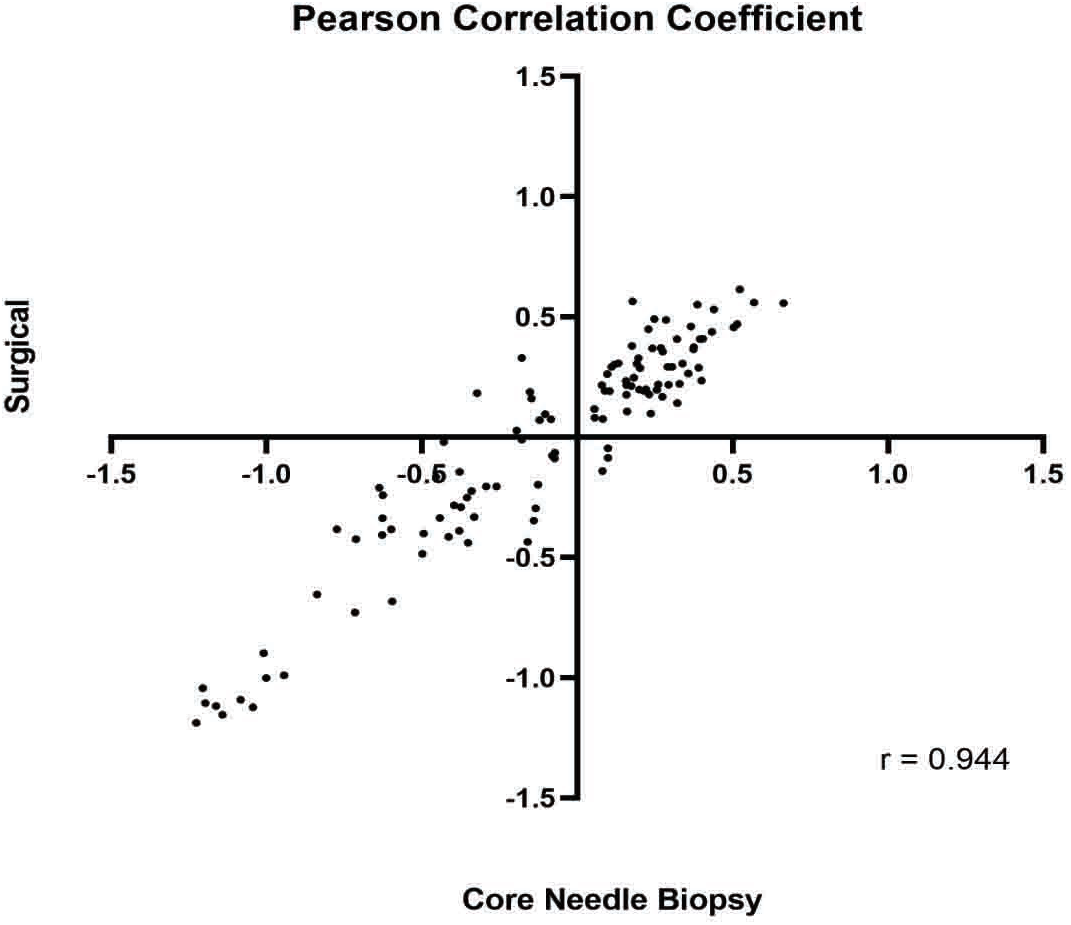
Correlation of MammaPrint Index between CNB and SR. MammaPrint index was determined for each matched CNB and SR tumor specimens (n = 113) and the correlation of matched samples was determined using Pearson correlation coefficient.

### Concordance of BluePrint molecular subtyping between CNB and SR tumor specimens

In a planned secondary analysis, the concordance of BluePrint molecular subtypes between the same matched CNB and SR samples was determined. CNB and SR tumors were in agreement for 98/99 (99.0% Luminal-type, 2/2 (100% HER2-type, and 11/12 (91.7% Basal-type tumors (**Table 3**. Overall, we determined the concordance of BluePrint between CNB and SR to be 98.2%. In addition, we observed a strong correlation of BluePrint index scores between CNB and SR specimens for Luminal-type (r = 0.92; p<0.001), HER2-type (r = 0.64; p<0.001), and Basal-type (r = 0.96; p<0.001) tumors (**Figure 2**).

**Table 3.** Concordance of BluePrint subtyping classification between CNB and SR

| BluePrint Result |  | SR |  |  | Total |
| --- | --- | --- | --- | --- | --- |
|  |  | Luminal-type | HER2-type | Basal-type |  |
| CNB | Luminal-type | 98 | 0 | 1 | 99 |
|  | HER2-type | 0 | 2 | 0 | 2 |
|  | Basal-type | 1 | 0 | 11 | 12 |
|  | Total | 99 | 2 | 12 | 113 |

**Figure 2.**
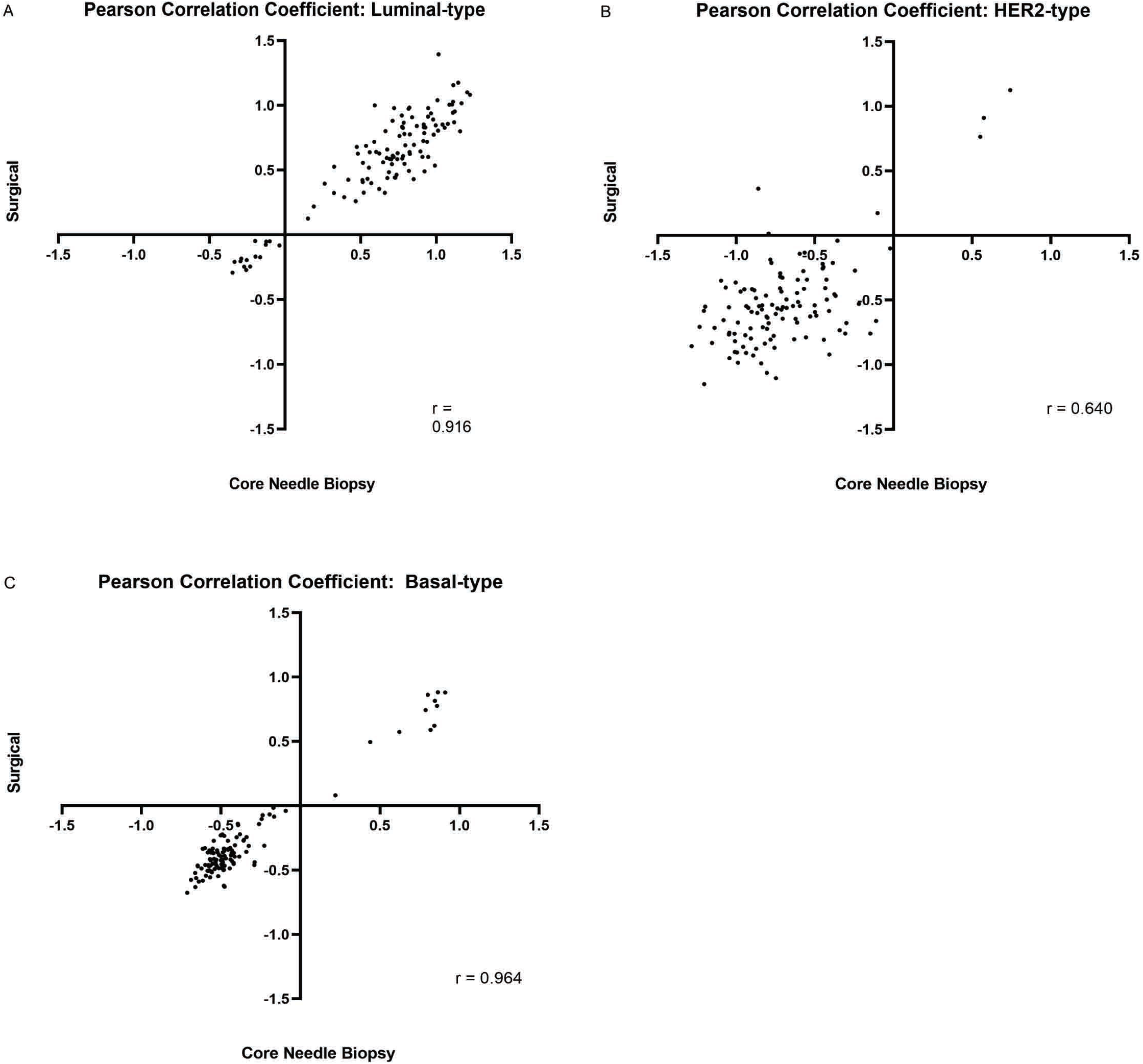
Correlation of BluePrint Index between CNB and SR. BluePrint generates three indices corresponding to Luminal-type, HER2-type, and Basal-type. Indices for (A) Luminal-type, (B) HER2-type, and (C) Basal-type were graphed and a Pearson correlation test was performed for each.

## Discussion/Conclusion

The utility for reliable prognostic information from preoperative core needle biopsies was established prior to the COVID-19 pandemic. However, as a part of the COVID-19 guidelines to conserve resources and triage patients, neoadjuvant therapy has predominantly been recommended for breast cancer patients. Thus, physicians rely on preoperative core biopsies to provide accurate subtyping and risk classification to guide therapy decisions. The traditional method for breast cancer classification and treatment is based on clinical features (tumor grade, receptor status, and Ki-67 as determined by IHC/FISH. Tumor grade is scored^19^ according to features such as tubule formation, mitotic rate, and nuclear pleomorphism. Multiple studies have analyzed the concordance of SR and CNB specimens in tumor grade, with overall agreement ranging from 64-77%^20-23^. In addition, numerous reports comparing SR and CNB samples in estrogen receptor (ER, progesterone receptor (PR, HER2, and Ki-67 IHC staining have been performed with overall agreement ranging from 84.0-99.1%, 77.9-94.3%, 80.0-98.8%, and 79.5-87.0% respectively, and a wide range of overall concordance (75-97%^22,24-31^.

Based on our previous performance and concordance studies^16,18,32^ using MammaPrint and BluePrint, we anticipated the overall agreement of MammaPrint results between CNB and SR tumor samples to be similar. As expected, we observed an overall agreement of 90.3%. These results confirm that MammaPrint risk classification from CNB are in high agreement with SR and provide consistent and accurate results. Importantly, discordant cases that are clinically relevant regarding how treatment decisions are affected are cases in which CNB is Low Risk and SR is High Risk, which account for less than 3% of patients in the current analysis. Thus, for 97% of patients in this study, treatment decisions and potential outcome are precise and consistent based on MammaPrint testing of the core needle biopsy.

In addition to our primary analysis, we determined the concordance of BluePrint genomic subtyping between the matched samples. Out of 113 tumors, 111 were in agreement resulting in an overall concordance of 98.2%, which is superior to previous reports of a concordance rate of 87.5% among intrinsic biological subtypes based on IHC assessment^22-24^. These results suggest that there is a clear benefit to knowing the activated pathways underlying a tumor’s biology through genomic testing compared to the conventional methods largely based on IHC. A limitation of our paired study is data maturity, in which patient follow-up data is currently unavailable to correlate outcome with MammaPrint and BluePrint results from CNB and SR samples. However, several studies, including the NBRST trial (NCT01479101), ISPY1 (NCT00033397), and ISPY2 (NCT01042379), have demonstrated accurate prediction in neoadjuvant treatment response and long-term outcome by MammaPrint and BluePrint on core needle biopsies^13,33-36^.

In summary, this analysis represents the largest study using prospective real-world data to evaluate the concordance of a genomic assay on matched CNB and SR samples. The high concordance rates of MammaPrint and BluePrint results between paired samples strongly support the utility of these assays to obtain reliable prognostic information on core biopsy tissue, which will expedite guide timely and appropriate treatment.

## Data Availability

The clinical datasets generated during and/or analyzed during the current study are available from the corresponding author on reasonable request. Raw array data have not been made publicly available as part of a collaboration agreement with the diagnostic company Agendia Inc.

## Acknowledgment

We would like to thank all of the patients who volunteered to participate in FLEX, as well as the FLEX site coordinators and investigaotrs.

## Author Contributions

All authors contributed to the study design and conceptualization. All authors contributed to collection and assembly of data. A.W., C.D., P.D., A.M., S.W., E.Y., W.A., and A.G., contributed to data analysis and interpretation. J.A.C., P.D., A.M., and S.W. contributed to manuscript preparation. All authors participated in the review and editing of the manuscript for publication.

## References

1. Ueda M, Martins R, Hendrie PC, et al. Managing Cancer Care During the COVID-19 Pandemic: Agility and Collaboration Toward a Common Goal. J Natl Compr Canc Netw. 2020:1–4.

2. Curigliano G, Cardoso MJ, Poortmans P, et al. Recommendations for triage, prioritization and treatment of breast cancer patients during the COVID-19 pandemic. Breast. 2020;52:8–16.

3. Surgeons ACo. COVID 19: Elective Case Triage Guidelines for Surgical Care. 2020.

4. Oncology SoS. Resource for Management Options of Breast Cancer During COVID-19. 2020.

5. Jakubowski DM, Bailey H, Abran J, et al. Molecular characterization of breast cancer needle core biopsy specimens by the 21-gene Breast Recurrence Score test. J Surg Oncol. 2020.

6. Muller BM, Brase JC, Haufe F, et al. Comparison of the RNA-based EndoPredict multigene test between core biopsies and corresponding surgical breast cancer sections. J Clin Pathol. 2012;65(7):660–662.

7. Lee J, Lee EH, Park HY, et al. Efficacy of an RNA-based multigene assay with core needle biopsy samples for risk evaluation in hormone-positive early breast cancer. BMC Cancer. 2019;19(1):388.

8. Van ‘t Veer L, Dal H, van de Vijver MJ, et al. Gene expression profiling predicts clinical outcome of breast cancer. Nature. 2002;415:6.

9. Cardoso F, van’t Veer LJ, Bogaerts J, et al. 70-Gene Signature as an Aid to Treatment Decisions in Early-Stage Breast Cancer. N Engl J Med. 2016;375(8):717–729.

10. Delahaye LJ, Wehkamp D, Floore AN, Bernards R, Van ‘t Veer L, Glas AM. Performance characteristics of the MammaPrint breast cancer diagnostic gene signature. Personalized Medicine. 2013;10(8):10.

11. Sapino A, Roepman P, Linn SC, et al. MammaPrint molecular diagnostics on formalin-fixed, paraffin-embedded tissue. J Mol Diagn. 2014;16(2):190–197.

12. Krijgsman O, Roepman P, Zwart W, et al. A diagnostic gene profile for molecular subtyping of breast cancer associated with treatment response. Breast Cancer Res Treat. 2012;133(1):37–47.

13. Whitworth P, Stork-Sloots L, de Snoo FA, et al. Chemosensitivity predicted by BluePrint 80-gene functional subtype and MammaPrint in the Prospective Neoadjuvant Breast Registry Symphony Trial (NBRST). Ann Surg Oncol. 2014;21(10):3261–3267.

14. Viale G, de Snoo FA, Slaets L, et al. Immunohistochemical versus molecular (BluePrint and MammaPrint) subtyping of breast carcinoma. Outcome results from the EORTC 10041/BIG 3-04 MINDACT trial. Breast Cancer Res Treat. 2018;167(1):123–131.

15. Nguyen B, Cusumano PG, Deck K, et al. Comparison of molecular subtyping with BluePrint, MammaPrint, and TargetPrint to local clinical subtyping in breast cancer patients. Ann Surg Oncol. 2012;19(10):3257–3263.

16. Mittempergher L, Delahaye LJ, Witteveen AT, et al. Performance Characteristics of the BluePrint(R) Breast Cancer Diagnostic Test. Transl Oncol. 2020;13(4):100756.

17. Glas AM, Floore A, Delahaye LJ, et al. Converting a breast cancer microarray signature into a high-throughput diagnostic test. BMC Genomics. 2006;7:278.

18. Mittempergher L, de Ronde JJ, Nieuwland M, et al. Gene expression profiles from formalin fixed paraffin embedded breast cancer tissue are largely comparable to fresh frozen matched tissue. PLoS One. 2011;6(2):e17163.

19. Elston CW, Ellis IO. Pathological prognostic factors in breast cancer. I. The value of histological grade in breast cancer: experience from a large study with long-term follow-up. Histopathology. 1991;19(5):403–410.

20. Connor CS, Tawfik OW, Joyce AJ, Davis MK, Mayo MS, Jewell WR. A comparison of prognostic tumor markers obtained on image-guided breast biopsies and final surgical specimens. Am J Surg. 2002;184(4):322–324.

21. Harris GC, Denley HE, Pinder SE, et al. Correlation of histologic prognostic factors in core biopsies and therapeutic excisions of invasive breast carcinoma. Am J Surg Path. 2003;27(1):4.

22. Burge CN, Chang HR, Apple SK. Do the histologic features and results of breast cancer biomarker studies differ between core biopsy and surgical excision specimens? Breast. 2006;15(2):167–172.

23. Daveau C, Baulies S, Lalloum M, et al. Histological grade concordance between diagnostic core biopsy and corresponding surgical specimen in HR-positive/HER2-negative breast carcinoma. Br J Cancer. 2014;110(9):2195–2200.

24. You K, Park S, Ryu JM, et al. Comparison of Core Needle Biopsy and Surgical Specimens in Determining Intrinsic Biological Subtypes of Breast Cancer with Immunohistochemistry. J Breast Cancer. 2017;20(3):297–303.

25. Lorgis V, Algros MP, Villanueva C, et al. Discordance in early breast cancer for tumour grade, estrogen receptor, progesteron receptors and human epidermal receptor-2 status between core needle biopsy and surgical excisional primary tumour. Breast. 2011;20(3):284–287.

26. Dekker TJ, Smit VT, Hooijer GK, et al. Reliability of core needle biopsy for determining ER and HER2 status in breast cancer. Ann Oncol. 2013;24(4):931–937.

27. Tamaki K, Sasano H, Ishida T, et al. Comparison of core needle biopsy (CNB) and surgical specimens for accurate preoperative evaluation of ER, PgR and HER2 status of breast cancer patients. Cancer Sci. 2010;101(9):2074–2079.

28. Arnedos M, Nerurkar A, Osin P, A’Hern R, Smith IE, Dowsett M. Discordance between core needle biopsy (CNB) and excisional biopsy (EB) for estrogen receptor (ER), progesterone receptor (PgR) and HER2 status in early breast cancer (EBC). Ann Oncol. 2009;20(12):1948–1952.

29. Mann GB, Fahey VD, Feleppa F, Buchanan MR. Reliance on hormone receptor assays of surgical specimens may compromise outcome in patients with breast cancer. J Clin Oncol. 2005;23(22):5148–5154.

30. Chen X, Sun L, Mao Y, et al. Preoperative core needle biopsy is accurate in determining molecular subtypes in invasive breast cancer. BMC Cancer. 2013;13:390.

31. Kombak FE, Sahin H, Mollamemisoglu H, et al. Concordance of immunohistochemistry between core needle biopsy and surgical resection of breast cancer. Turk J Med Sci. 2017;47(6):1791–1796.

32. Beumer I, Witteveen A, Delahaye L, et al. Equivalence of MammaPrint array types in clinical trials and diagnostics. Breast Cancer Res Treat. 2016;156(2):279–287.

33. Whitworth P, Pellicane JV, Baron P, et al. Abstract PD9-01: 5-year outcomes in the NBRST trial: Preoperative MammaPrint and BluePrint breast cancer subtype is associated with neoadjuvant treatment response and survival. Cancer Research. 2021;81(4 Supplement):PD9-01-PD09-01.

34. Gluck S, de Snoo F, Peeters J, Stork-Sloots L, Somlo G. Molecular subtyping of early-stage breast cancer identifies a group of patients who do not benefit from neoadjuvant chemotherapy. Breast Cancer Res Treat. 2013;139(3):759–767.

35. Esserman LJ, Berry DA, Cheang MC, et al. Chemotherapy response and recurrence-free survival in neoadjuvant breast cancer depends on biomarker profiles: results from the I-SPY 1 TRIAL (CALGB 150007/150012; ACRIN 6657). Breast Cancer Res Treat. 2012;132(3):1049–1062.

36. Wolf DM, Yau C, Sanil A, et al. DNA repair deficiency biomarkers and the 70-gene ultra-high risk signature as predictors of veliparib/carboplatin response in the I-SPY 2 breast cancer trial. NPJ Breast Cancer. 2017;3:31.

